# Dynamic AI-assisted Ipsilateral Tissue Matching for Digital Breast Tomosynthesis

**DOI:** 10.1101/2024.12.02.24318238

**Authors:** Stephen Morrell, Michael Hutel, Oeslle de Lucena, Cristina Alfaro Vergara, Georgiana Zamfir, Charlottefreya Longman, Rumana Rahim, Sophia O’Brien, Elizabeth S. McDonald, Samantha Zuckerman, John Scheel, Anna Metafa, Nisha Sharma, Sebastien Ourselin, Jorge Cardoso, Juliet Morel, Keshthra Satchithananda, Emily Conant

## Abstract

**Structured Abstract:** *Purpose:* To compare Digital Breast Tomosynthesis (DBT) tissue matching errors with and without artificial intelligence (AI) assistance to typical screen-detected breast tumor sizes, evaluating whether AI ameliorates lesion mislocalization beyond tumor boundaries, especially for nonexpert radiologists. The technology category is deep learning.

*Materials and Methods:* This multicenter retrospective feasibility study conducted in April 2022 – July 2023 included 12 radiologists (mean age, 42 years ± 8) interpreting 94 lesion regions of interest in 30 women. Readers performed annotations with and without AI assistance after a minimum four-week washout period. The root mean square errors (RMSE) and maximum distance errors (MDE) were measured relative to consensus references. Stratifications included radiologist expertise (*≥* 5 vs *<* 5 years), lesion abnormality, and AI warnings. The Wilcoxon signed-rank test was used to assess statistical significance.

*Results:* Across all abnormal lesions, mean RMSE was 32% higher without AI (11.70*mm* vs 8.88*mm*, *p* = .049), and mean maximum distance errors were 37.5% higher (20.68*mm* vs 15.08*mm*, *p* = .036). Non-expert radiologists showed the largest benefit: for abnormal lesions without AI warnings, RMSE was 61.9% higher without AI (12.20*mm* vs 7.57*mm*, *p* = .010) and maximum distance error was 67.5% higher (15.76*mm* vs 9.47*mm*, *p* = .028). These reductions are clinically relevant given typical screen-detected breast tumor sizes (median, 13*mm* [IQR: 9–20]).

*Conclusion:* AI-assisted tissue matching significantly reduced DBT localization errors, particularly for non-experts handling challenging cases. By keeping errors below typical tumor dimensions, AI may improve diagnostic precision and reduce risks of missed or mischaracterized lesions.

**Summary Statement:** Dynamic artificial intelligence assisted tissue matching in digital breast tomosynthesis improves localization accuracy for non-expert radiologists, with errors in abnormal cases significantly larger (67.5%, *p ≤* 0.05) without assistance.

**Key Points:** In this multicenter retrospective study of 94 Regions of Interest (ROI) analyzed by 12 radiologists across 5 hospitals, manual tissue matching was found to have errors 32% higher in abnormal cases than AI-assisted tissue matching (*p <* 0.05).

For non-expert radiologists interpreting abnormal cases (excluding cases with AI warnings) without AI assistance, the root mean squared distance errors (RMSE) were found to be 61.9% higher (12.20*mm* vs 7.57*mm*, *p <* 0.01) and the maximum distance errors (MDE) was 67.6% higher (15.76*mm* vs 9.47*mm*, *p <* 0.05) than when using AI assistance.

For challenging cases, many non-expert readers’ MDE without AI assistance (75th percentile: 20.21*mm*) exceeded the largest tumor dimensions (75th percentile: 20*mm*), while AI-assisted errors (75th percentile: 11.94*mm*, *p <* 0.05) remained within median tumor sizes (12*mm*), potentially preventing correlation with non-lesion tissue.

## 1 Introduction

Multiple view mammography is a well-established standard, with studies showing that two-view mammography results in a 15-20% reduction in interval cancer rates[1], and that cancers detected using two views were often smaller and had less aggressive characteristics than those missed by single view[2]. However, this advantage relies on accurate correlation between views for localizing, classifying, and confirming lesions. Mismatches can result in missed cancers, unnecessary imaging and biopsies, incorrectly targeted biopsies leading to false negatives, and reduced radiologist confidence, and may negate these benefits[3].

Digital Breast Tomosynthesis (DBT) offers substantial improvements in breast cancer detection over planar mammography, with a 30%–40% higher detection rate[4, 5]. However, DBT interpretation requires radiologists to mentally correlate tissue across different non-orthogonal projection angles, and between 60-80 depth slices per view. This complexity adds cognitive load, doubles interpretation times, and imposes specialized training requirements[3, 6]. Scroll bars partially ameliorate this, but lack precision and mental effort persists in translation between views[3]. When combined with unpredictable tissue deformation and change in appearance between views[7], these factors create a fundamentally more demanding interpretation environment.

Current computer-aided detection (CAD) systems, while promising in cancer detection[8, 9], do not address the fundamental challenge of dynamic tissue matching between views[10, 11], do not facilitate interrogation of Regions of Interest (ROIs), and are not yet capable of replacing human readers in double-reading workflows[9, 12].

The likelihood of mislocalization remains substantial Despite DBT’s proven benefits. A pilot study noted a 23% error rate in mental tissue matching even among experienced breast radiologists. Furthermore, a 2021 case in our institution progressed from Stage 1 to Stage 4 after three specialists incorrectly matched suspicious densities between views (Appendix S1). Such errors may be more frequent outside specialized centers; approximately 70% of all mammographic screening in the United States is performed by non-specialist radiologists [13], with less exposure to DBT correlation.

We hypothesize that a dedicated AI system to facilitate ipsilateral tissue matching could improve reader confidence and localization accuracy, particularly for non-expert radiologists. By maintaining accuracy within or below the 13*mm* median diameter of screen-detected breast tumors[14], AI assistance could improve efficiency and help minimize the risk of correlating suspicious findings with non-lesion tissue. Therefore, the purpose of this study was to compare DBT tissue matching errors with and without AI assistance to typical early-stage tumor sizes, assessing whether AI can help prevent significant mislocalization, especially for non-expert readers.

## 2 Materials and Methods

### 2.1 Study Approval and Ethics

We conducted a retrospective two-part, multicenter feasibility study including multiple UK and USA sites and the from April 2022 to December 2023. All cases were complete unilateral assessment cases comprising craniocaudal (CC) and mediolateral oblique (MLO) views of DBT and co-registered full-field digital mammography (FFDM) images from Hologic devices (Hologic, Inc., Marlborough, Mass), and excluding scout views, post-biopsy images, synthetic 2D reconstructions (C-View) images, cases with multiple DBT assessments and cases. The study comprised two distinct experiments; a subjective evaluation of the reading tool on disparate data and object localization accuracy on common data. The study was approved by KCH’s Electronic Records Research Initiative (KERRI), with a waiver of informed consent and data were fully anonymized before analysis. Use of the OPTIMAM dataset was permitted by license[15].

### 2.2 Medical Device Description

ViewFinder is an FDA-cleared (K223501) DBT reading tool [16] addressing the tissue-matching challenge. Unlike traditional CAD systems, ViewFinder enhances interpretation by mapping corresponding tissue between views using tissue properties and appearance. This enhances ipsilateral DBT pair reading through three key features. First, it provides dynamic matching - as a user moves their cursor in a source tomosynthesis view, the correlated 3D location in the target view is highlighted by automatically scrolling to the correct slice and plotting a target ellipse containing the matching tissue. The ellipse size varies with matching confidence, with a minimum size of 35*mm* × 50*mm*, and a warning is shown if mapping uncertainty is high. Second, it displays reference measurements, showing in the target both a line equidistant from the pectoral muscle and an arc equidistant from the nipple relative to the cursor position in the source view. Third, it provides coronal views in both compressed and uncompressed states for frontal localization (Appendix S2). These multiple synchronized views and precise measurements supersede traditional scroll bars, quadrant and clock face position descriptions, providing reproducible spatial coordinates across all views. This enhanced precision is particularly valuable in remote diagnostic workflows, where accurate communication between interpreting radiologists and on-site technologists performing additional imaging is crucial, and for biopsy and surgical planning where exact lesion localization is essential.

### 2.3 Part 1: Subjective Evaluation on Varying Assessment Data

The first part was a subjective evaluation of the AI system’s usefulness by 14 dedicated breast radiologists across 5 international sites: King’s College Hospital NHS Foundation Trust, London, UK, Princess Royal University Hospital, Orpington, UK, Leeds Teaching Hospitals NHS Trust, Leeds, UK, Department of Radiology, Vanderbilt University Medical Center, Nashville, TN, USA and the Department of Radiology, Hospital of the University of Pennsylvania, Philadelphia, PA, USA. They were stratified by post-training breast imaging experience into nine “experts” (*≥* 5 years) and five “non-experts” (*<* 5 years), following [17], which shows significant performance maturation within five years. In the US, this typically reflects residency plus a breast fellowship, whereas in the UK it involves specialty training plus optional post-Certificate of Completion of Training (CCT) fellowships. All participants practiced exclusively in breast imaging; no general radiologists were included.

#### 2.3.1 Dataset

For Part 1, data were sourced from two sets for variety and permission reasons. For KCH and PRUH, 11 unilateral cases were randomly selected with permission from KCH’s 2020-2021 screening assessments which were at least six months prior to ensure adequate washout from the radiologist’s hospital experience and de-identified by KCH. To ensure an unbiased evaluation of the AI system’s performance and to prevent data leakage, all cases previously used for training and testing the AI model were excluded from the Parts 1 and 2 datasets. For centers other than KCH and PRUH, 11 cases acquired between 2010 and 2021 were randomly selected from previously de-identified OPTIMAM data[15].

#### 2.3.2 Evaluation Metrics

Radiologists were blinded to patient metadata and followed a prescribed hanging protocol; first reviewing the 2D pair, then the DBT pair, with freedom to revert to any combination as needed. For each case, four key metrics were recorded: whether objects in the ipsilateral view fell within the AI system’s predicted oval, if AI assistance increased confidence in tissue localization and matching between projections, whether the case was too obvious for the AI system to be useful, and a subjective score of its usefulness on a 0-10 scale (where 0 indicated ”hindered reading”, 5 meant neither helped nor hindered, and 10 signified helped reading).

Following each reading session, radiologists completed a standardized questionnaire that evaluated their overall confidence correlating lesions with ViewFinder on a 0-10 scale (0 indicating reduced confidence, 5 no change, and 10 maximum confidence improvement), recording their levels of experience, general assessments of the AI system’s usefulness, its impact on reading speed and tissue matching accuracy, and potential influence on recall rates.

Statistical significance for case-level subjective usefulness scores was assessed using a linear mixed-effects model to account for the nested structure of multiple readings per radiologist and multiple radiologists per breast, with random effects for both radiologists and breasts. For radiologist-level confidence scores, which had no nesting structure (one score per radiologist), significance was assessed using a one-sample t-test against the neutral score of 5, with power analysis to validate sample size adequacy.

### 2.4 Part 2: Multi-site Objective Localisation Performance on Common Assessment Data

The second part analyzed the inter-reader variability of matching tissue location in an assessment setting. The same set of radiologists as in Part 1 excluding two from VUMC interpreted all Part 2 cases (Section 2.4.1). For each case, readers reviewed the FFDM images then by the DBT images, observing standard clinical protocols. Readers marked all tissue potentially influencing recall decisions on the DBT images, including both suspicious findings and tissues that would be inspected but ultimately dismissed. Readers drew tight bounding boxes around these features on the most representative DBT slice in both projections (CC and MLO). Each reader completed two interpretation sessions, separated by a minimum four-week washout period to minimize recall bias. ViewFinder availability was systematically alternated between sessions and cases using a balanced crossover design. Readers were blinded to case outcomes, prior imaging, clinical history, and other readers’ interpretations throughout the study. All reading sessions were timed and conducted in the respective departments of breast radiology.

#### 2.4.1 Dataset

Under standard assumptions, a study would require a sample size of 27 (number of paired samples) to achieve a power of 80% and a level of significance of 5% (one sided), for detecting a mean error reduction of 25% between pairs, assuming a median tumor size of 13*mm* diameter and a expected variance equal to the median radius (6.5 mm) [18, 19] which aligns with the reported 5 mm threshold, beyond which localization errors significantly increase the risk of failed surgical removal, supporting the assumption that variance should be at least comparable to this critical threshold[20]. Due to the conservative nature of the error estimate above, a decision was made to overpower the study by a factor of 3 to further minimize the risk of rejecting the null hypothesis while being constrained by the methodological demands of reference ROI annotation. This overpowering of the sample size was later validated by a more recent Ultrasound localisation study with 19.9mm error and Std 16.1mm [21], demonstrating that a 25% reduction in error would require a sample size of 67.

For Part 2, 10 cases without biopsy (normal), and 20 cases with wide bore needle (WBN) biopsy-proven findings were randomly selected with sample sizes set by two consultant radiologists with special interest in breast imaging. Of the 20 cases, 4 (20%) were benign and 16 (80%) malignant, similar proportions to OPTIMAM with 27% and 73% respectively, with a broad selection of clinical outcomes. Density, ethnic and BI-RADS data were not available for either dataset. Refer to Appendix S3 for reader and data characteristics.

#### 2.4.2 Reference Standard Annotations

To generate reference standard annotations, we used a consensus derived from all 12 radiologists’ annotations, considering both ViewFinder (VF) ON and VF OFF, without revealing their sources. Location information was not available in the metadata. The consensus process involved two experienced breast radiologists independently reviewing the 12 readers’ annotations to establish reference locations using Weasis[22]. This approach acknowledges that no absolute ground truth exists for tissue matching in DBT, making multiple expert consensus the most reliable available standard. Initially, all radiologist annotations were aligned to the mid-slice of the DBTs. The region with the highest consensus among the annotations was selected as the initial estimate of the reference standard. Afterwards, a second independent clinician, not involved in the initial annotation process described in Section 2.4, reviewed and amended the reference standard annotation’s position and slice for validation. All changes were actively discussed between the two experts with qualifications: MSc Radiologic Sciences, MSc Health Sciences Education, PhD candidate in Biological and Medical Engineering, and Medical Technologist specializing in mammography; and MBBS, FRCR, Consultant Radiologist with a special interest in breast imaging, and Director of Screening, respectively. This resulted in 94 lesions in Part 2.

#### 2.4.3 Evaluation Metrics

We evaluated the location accuracy of radiologist annotations computed with VF on versus VF off. For each case, a source view and a target view (i.e., the other view CC vs. MLO) were initially selected. Reference standard ROIs were determined in the source view using the Intersection over Union (IoU) criterion. Each radiologist annotation was assigned to the reference standard ROI with the highest IoU; annotations without a match were discarded.

Next, we calculated the root mean squared error (RMSE) of the radiologist annotations relative to their respective reference standard ROIs, as well as the maximum distance of a radiologist annotation from its corresponding reference standard ROI in the source view and its counterpart in the ipsilateral (target) view. The metrics were then recomputed by swapping the source and target views, and the final result was obtained by averaging the two sets of measurements. To assess the statistical significance of the RMSE and maximum distance, we used the Wilcoxon signed-rank test [23].

## 3 Results

### 3.1 Part 1: Multi-site Subjective Evaluation

In 126 breast readings, 12 radiologists scored the AI system *≈* 6.21 subjective usefulness on average on a 10-point scale, significantly higher than the neutral score of 5 when accounting for repeated measures (mixed-effects model, *p ≤* 0.001). 42.1% of readings (53/126) received scores of 7 or higher, predominantly in non-obvious cases where tissue matching was more challenging. Lower scores (5-6) typically corresponded to cases with easily visible large masses spanning multiple slices in both views (Fig. 2).

**Figure 1:**
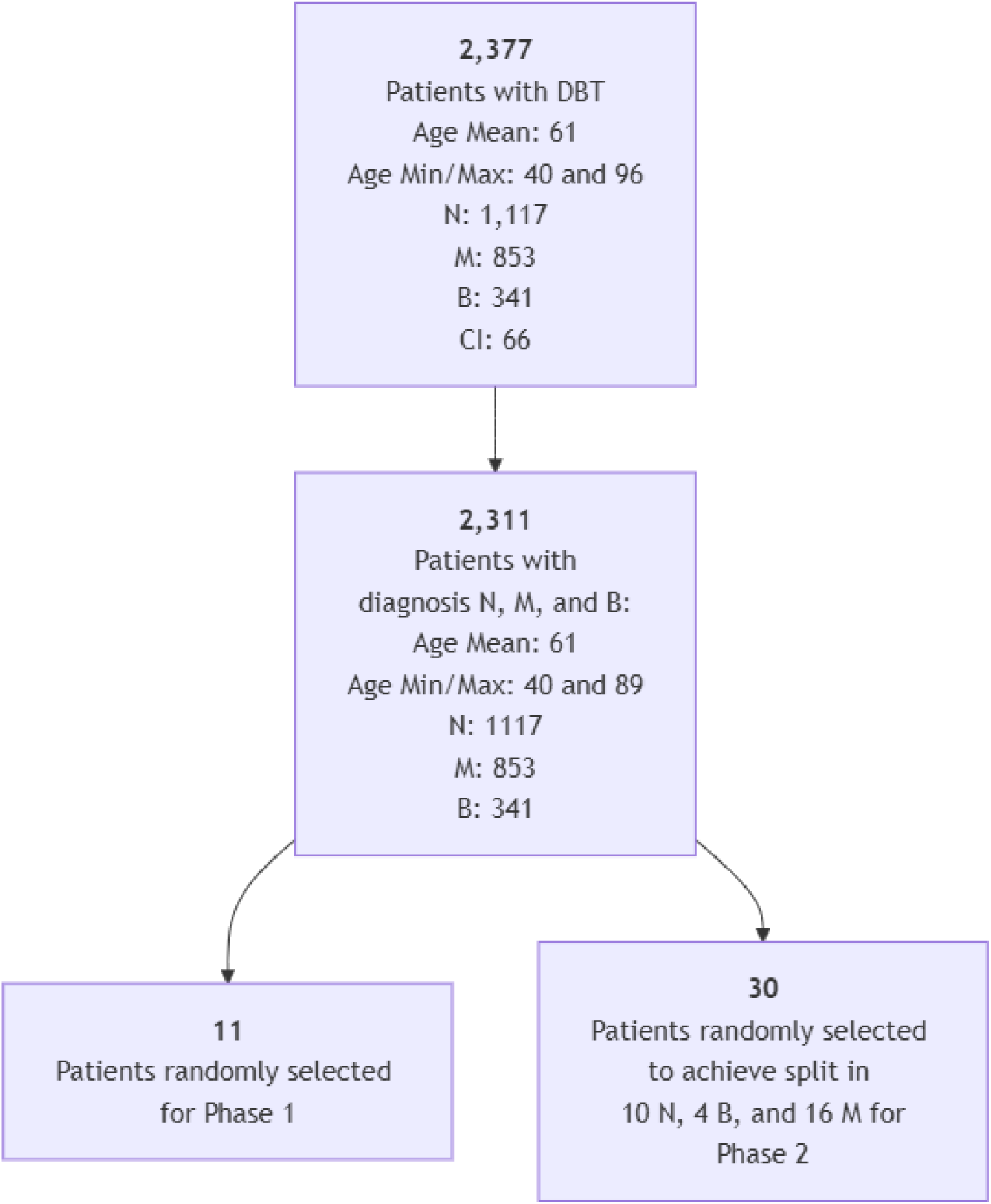
Part 2 case selection STARD diagram for OPTIMAM[15] cases.

**Figure 2:**
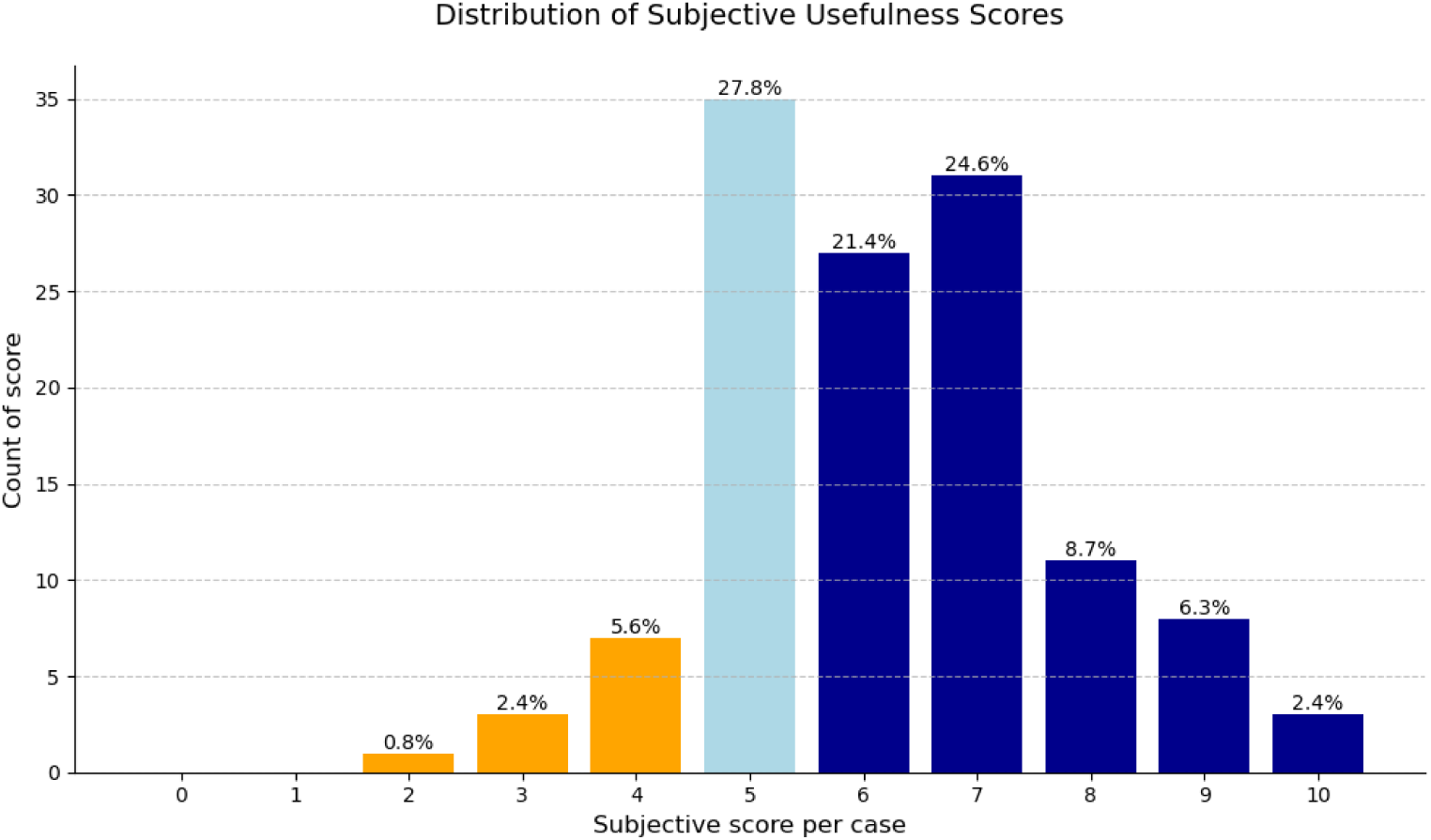
Distribution of radiologist-reported usefulness scores for ViewFinder across 126 breast readings (mean score = 6.21, *p ≤* 0.001 compared to neutral score of 5, mixed-effects model). Scores of 0-4 indicated ViewFinder hindered reading (8.7% of ratings) (orange), 5 indicated neither help nor hindrance (light blue), and 6-10 indicated ViewFinder helped reading (63.5% of ratings) (dark blue). 42.1% of readings received scores of 7 or higher.

In post-study evaluations, radiologists reported high confidence gains in localization, with a mean confidence score of 7.03 (one-sample t-test, *p <* 0.001, Cohen’s *d* = 1.48). The study achieved *>* 99% statistical power with 16 radiologists. 81.25% of radiologists reported increased confidence (scores 6-10), with 50% reporting a score of 7, and 18.75% reporting high confidence scores (9-10). The remaining 18.75% reported no change in confidence (score 5), while no radiologists reported decreased confidence (Fig. 3).

**Figure 3:**
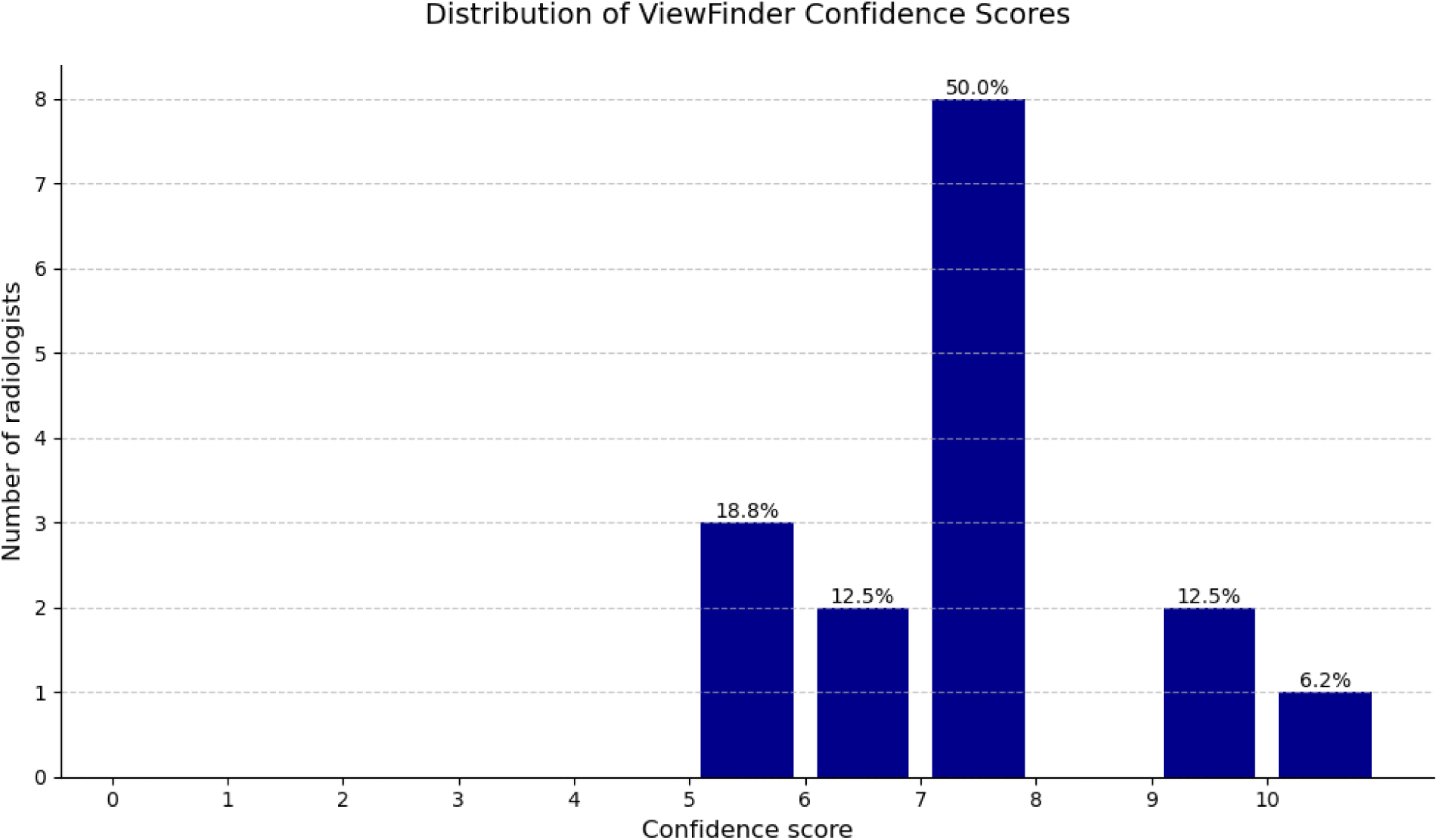
Distribution of radiologist-reported confidence scores for ViewFinder (n=16 radiologists). Radiologists rated ViewFinder’s effect on their confidence in correlating lesions on a scale where 0 indicated reduced confidence, 5 indicated no change, and 10 indicated maximum confidence improvement. The mean score was 7.03, with 81.25% of radiologists reporting increased confidence (scores 6-10). Notably, 50% of radiologists reported a score of 7, and 18.75% reported high confidence scores (9-10). The remaining 18.75% reported no change in confidence (score 5), while no radiologists reported decreased confidence (scores below 5).

### 3.2 Part 2: Multi-site Objective Localisation Performance on Common Assessment Data

Twelve radiologists annotated 30 patients resulting in 94 ROIs meeting criteria, and 710 paired data points for localization evaluation in CC and MLO. We evaluated the accuracy of these annotations (Section 2.4) against the reference standard annotations (Section 2.4.2) using the metrics outlined in Section 2.4.3 under the following stratification scenarios: (1) all reference standard annotations (All), (2) all reference standard annotations excluding normal cases (Abnormal), (3) all reference standard annotations excluding normal and warning cases (Abnormal, No Warnings), (4) all reference standard annotations excluding normal, warning, and non-expert-labelled cases (Abnormal, No Warnings, Experts), and (5) all reference standard annotations excluding normal, warning, and expert-labelled cases (Abnormal, No Warnings, Non-experts). Table **??** presents the leave-one-out cross-validation results for RMSE and maximum distances across all stratifications. Figure 4 illustrates the distribution of individual data points using violin plots. Analysis of reading times showed no statistically significant difference between readings with and without AI assistance.

**Figure 4:**
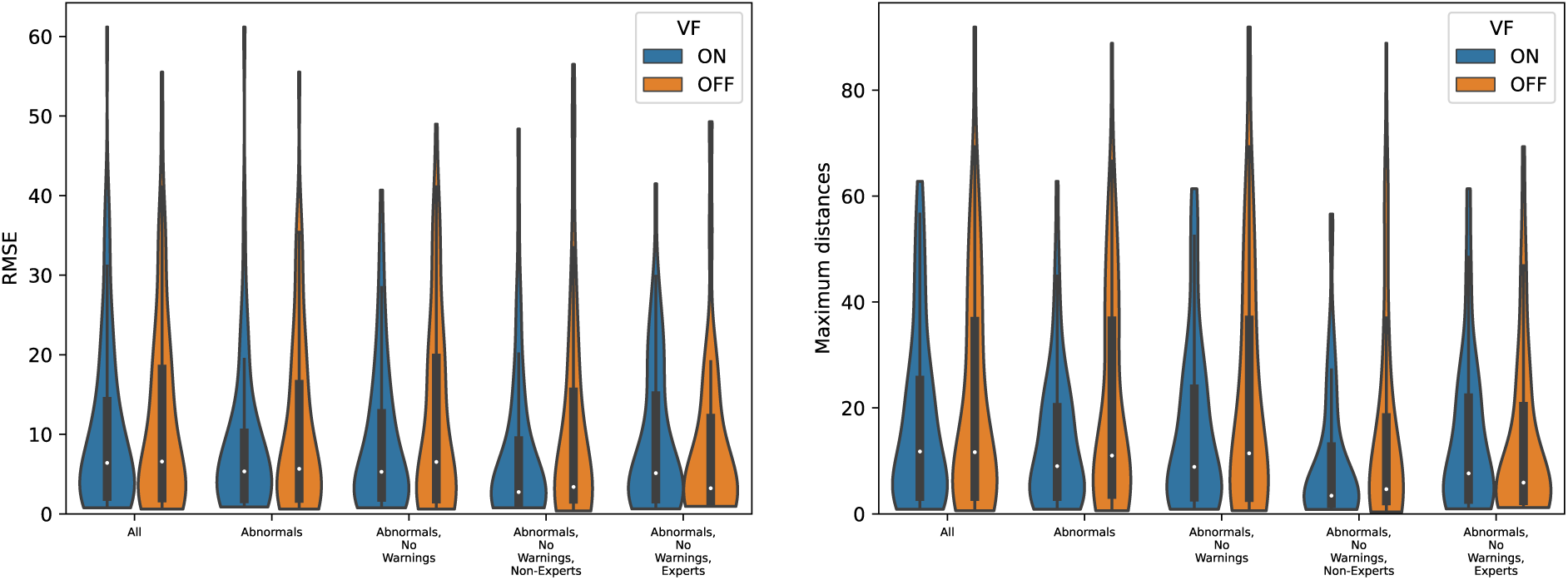
Violin distribution plots for RMSE (left) and Maximum Distance (right) relative to reference standard mappings across different stratifications, comparing results with (ON) and without (OFF) ViewFinder. A notably lower error is observed when ViewFinder is used, especially in cases involving abnormal subgroups, cases without warnings, and non-expert users (RMSE: *p <* 0.01, Max: *p <* 0.05.)

**Figure 5:**
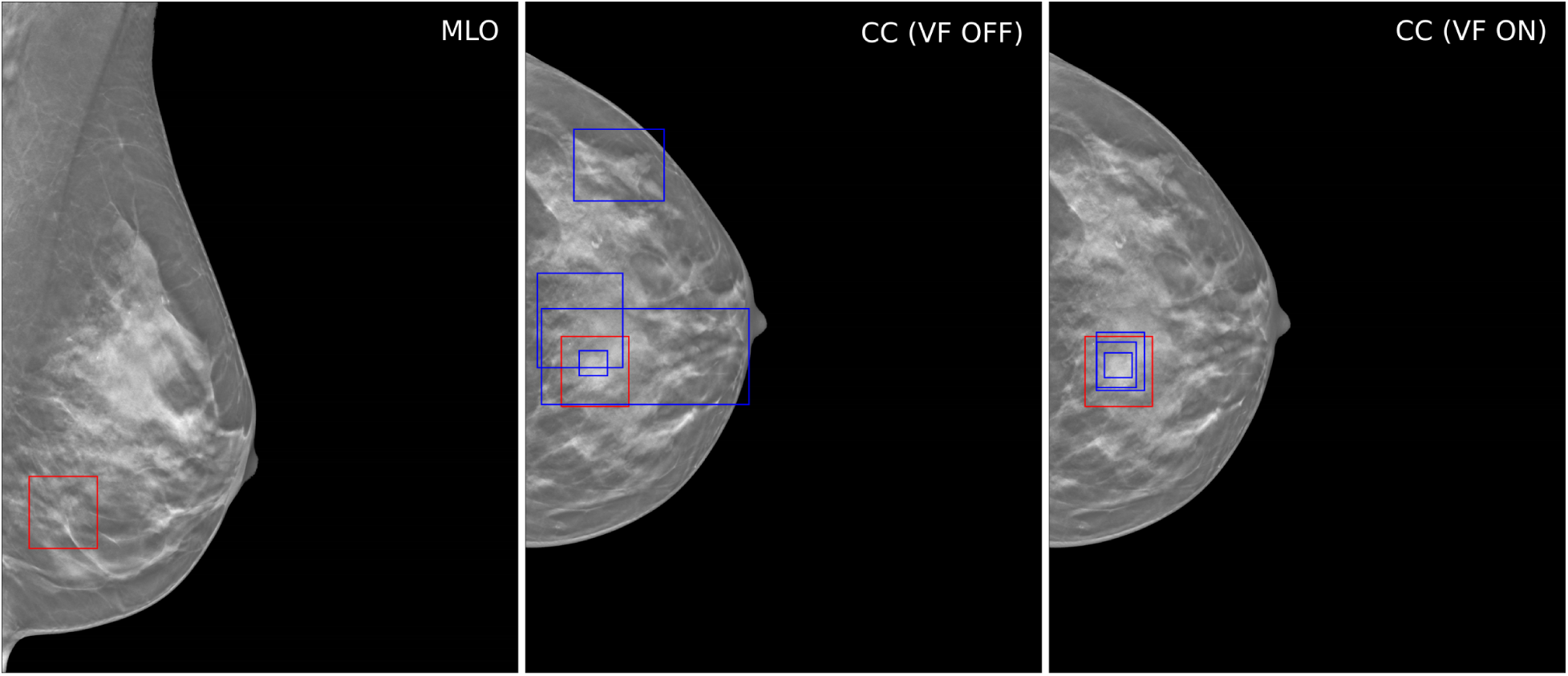
Example case demonstrates improved accuracy in lesion location with VF on versus VF off. MLO view shows the source reference-standard location (in red) while the CC images show a series of radiologist-mapped locations (blue) both VF ON and VF OFF compared to the reference standard location (in red).

## 4 Discussion

Our study demonstrates that dynamic AI-assisted tissue matching significantly improves localization accuracy in DBT interpretation. The data shows that mental matching errors without AI assistance were 32% larger (p = 0.049) in abnormal cases, with maximum distance errors 37.5% larger (p = 0.036) when not using AI assistance. This improvement was most pronounced in challenging cases (90th percentile), where maximum distance errors were reduced from 48.12*mm* to 26.21*mm* for non-expert readers. The clinical significance emerges when considering typical early-stage breast cancer dimensions. With the median screen-detected size of 13*mm*[14], T1 *≤* 20*mm* and T2 tumors *≤* 50*mm*[19, 18], our observed reduction in matching errors represents a meaningful improvement in clinical practice. When tissue matching errors exceed tumor dimensions—as observed in the control group with maximum errors of 48.12*mm* —there is increased risk of correlating suspicious areas with non-lesion tissue, potentially leading to mischaracterizations, unnecessary imaging, incorrectly targeted biopsies, or missed diagnoses. The most notable improvement occurred among non-expert radiologists interpreting abnormal cases without warnings, where mental matching showed 61.9% larger RMSE (12.20*mm* vs. 7.57*mm*, p = 0.010) and 67.5% larger maximum distance errors (15.76*mm* vs. 9.47*mm*, p = 0.028). This improvement is particularly relevant given that 70% of USA mammograms are interpreted by non-specialist radiologists, who may represent a key beneficiary group for this technology. The magnitude of mental matching errors in the harder cases (90th percentile) was 35.48*mm* vs. 20.66*mm* with AI assistance in mean error between raters. Expert radiologists showed minimal accuracy improvement with AI assistance (RMSE change of 0.6%, p = 0.91), suggesting they have developed robust mental models through years of practice. However, they reported increased confidence and reduced mental effort when using the tool, indicating benefits beyond pure accuracy metrics. This aligns with our subjective usefulness findings, where the AI system achieved an average score of 6.21 on a 10-point scale, with 42.1% of readings receiving scores of 7 or higher. Our results complement existing computer-aided detection (CAD) systems for DBT. While traditional CAD primarily focuses on highlighting suspicious regions and has shown limited success in improving diagnostic accuracy, our approach addresses the fundamental challenge of tissue matching between views. The 23% error rate in manual tissue matching observed in our pilot study aligns with previous literature on DBT interpretation challenges and highlights the need for targeted solutions. The findings suggest several potential benefits for clinical practice. Improved tissue matching accuracy could reduce satisfaction of search errors, where radiologists prematurely stop searching after finding initial suspicious areas. The synchronized views may help counter this tendency by enabling systematic verification across views. Additionally, the system’s ability to communicate precise lesion locations could improve workflow efficiency for the entire care team, including imaging technologists and surgeons[20].

Several limitations warrant consideration. Our evaluation metrics focused on spatial accuracy rather than diagnostic outcomes, and the relatively small dataset (94 labelled ROIs, 12 readers, driven by burdensome reference ROI determination and data processes) suggests the need for larger validation studies. The preponderance of malignant cases may have prompted increased reader attention compared to clinical practice. The AI model’s performance varies with case complexity and tissue characteristics, necessitating explicit warnings in challenging cases. Our findings on reading efficiency were inconclusive and require further investigation with extended training periods. Future research directions should include prospective studies evaluating impact on cancer detection rates and false positive recalls, investigation of potential synergies with existing CAD systems, and assessment of the technology’s role in radiologist training and quality improvement programs. Additionally, studies should evaluate the impact of standardized spatial communication on downstream clinical workflows, including ultrasound localization, additional view positioning, and surgical planning precision.

Dynamic AI-assisted tissue matching significantly improves localization accuracy in DBT interpretation, particularly for non-expert radiologists analyzing challenging cases. The reduction in matching errors to clinically relevant dimensions, combined with increased reader confidence, suggests this technology could enhance breast cancer detection accuracy. While questions remain about long-term clinical impact and implementation strategies, the proposed technology has the potential to impact in breast cancer screening and improving patient outcomes.

## Ethics Statement

This study was conducted under ethical approval from the KCH’s Electronic Records Research Interface (KERRI) of King’s College Hospital NHS Foundation Trust, London, UK. Additional images were accessed from the Optimisation of Breast Cancer Detection using Mammography (OPTIMAM) database at the Royal Surrey NHS Foundation Trust under contractual license in accordance with the database’s research use terms. All data collection and analysis were performed in accordance with relevant guidelines and regulations. All data were anonymized prior to analysis.

## Data Availability

All data produced in the present work are contained in the manuscript

https://www.cancerresearchhorizons.com/our-portfolio/our-licensing-opportunities/optimam-mammography-image-database-omi-db

## Acknowledgments

Some of the images and data used in this publication are derived from the OPTIMAM imaging database [15], we would like to acknowledge the OPTIMAM project team and staff at the Royal Surrey NHS Foundation Trust who developed the OPTIMAM database, in particular Mark Halling-Brown, PhD, for his leadership and dedication in assembling the exemplary dataset, and Cancer Research Horizons who funded the database. CAV thanks the Chilean National Agency for Research and Development (ANID) for the National PhD Grant (No. 21212497) and the Universidad de Tarapacá, Arica, Chile (DEC.EX. No. 00.0.587/2020) for their support. Additionally, we acknowledge King’s College Hospital NHS Foundation Trust, London, UK for providing anonymized imaging data used in this study and for their continued support in advancing breast imaging research.

---

**Table 1:** Leave-one-out cross validation statistics for the RMSE and Maximum distances for all gold standard annotations for different scenarios. All statistics are bootstrapped using a leave-one-out cross validation. Statistical significance is marked with (*).

| Stratification | Statistic | RMSE |  |  |  | Maximum Distance |  |  |  |
| --- | --- | --- | --- | --- | --- | --- | --- | --- | --- |
|  |  | VF ON<br>mean(std) | VF OFF<br>mean(std) | %Rel. Change<br>mean(std) | p-value | VF ON<br>mean(std) | VF OFF<br>mean(std) | %Rel. Change<br>mean(std) | p-value |
| 1. All | Mean | 10.14 (0.27) | 11.42 (0.33) | 12.72 (3.32) | 0.87 | 16.57 (0.57) | 20.23 (0.59) | 22.14 (4.01) | 0.06 |
|  | Median | 6.32 (0.32) | 6.44 (0.31) | 2.17 (5.36) |  | 10.49 (0.91) | 10.83 (0.37) | 3.93 (8.66) |  |
|  | 75th Percent | 13.78 (0.68) | 17.65 (0.77) | 28.41 (9.50) |  | 24.38 (1.00) | 34.61 (2.33) | 42.13 (10.14) |  |
|  | 90th Percent | 24.67 (0.40) | 29.42 (1.35) | 19.29 (5.85) |  | 42.45 (1.99) | 50.93 (0.90) | 20.23 (5.55) |  |
| 2. Abnormals | Mean | 8.48 (0.16) | 10.57 (0.37) | 24.62 (3.77) | 0.37 | 13.60 (0.38) | 19.89 (0.68) | 46.26 (4.66) | <0.05* |
|  | Median | 4.98 (0.65) | 5.57 (0.45) | 13.12 (11.58) |  | 8.46 (0.79) | 10.33 (0.67) | 22.88 (11.09) |  |
|  | 75th Percent | 10.34 (0.54) | 16.25 (0.71) | 57.62 (10.70) |  | 19.90 (0.51) | 36.20 (0.62) | 82.04 (5.90) |  |
|  | 90th Percent | 19.88 (0.47) | 27.58 (2.49) | 38.68 (10.98) |  | 30.17 (0.58) | 50.60 (0.71) | 67.78 (3.39) |  |
| 3. Abnormals,<br>No Warnings | Mean | 8.88 (0.46) | 11.70 (0.45) | 32.06 (5.85) | <0.05* | 15.08 (0.91) | 20.68 (0.86) | 37.52 (8.30) | <0.05* |
|  | Median | 4.93 (0.47) | 6.22 (0.47) | 27.06 (12.89) |  | 8.16 (0.85) | 10.76 (0.45) | 33.30 (15.35) |  |
|  | 75th Percent | 12.81 (0.90) | 18.98 (0.61) | 48.79 (9.80) |  | 22.78 (1.30) | 32.79 (2.99) | 44.06 (11.17) |  |
|  | 90th Percent | 21.87 (1.49) | 30.95 (2.20) | 41.87 (9.94) |  | 38.11 (3.35) | 52.97 (2.11) | 40.15 (14.78) |  |
| 4. Abnormals,<br>No Warnings,<br>Experts | Mean | 8.77 (0.71) | 8.81 (0.59) | 0.58 (3.93) | 0.91 | 13.24 (1.12) | 12.75 (0.78) | -3.42 (3.42) | 0.61 |
|  | Median | 4.78 (0.47) | 3.40 (0.19) | -28.37 (6.26) |  | 6.95 (0.51) | 5.65 (0.43) | -18.59 (4.48) |  |
|  | 75th Percent | 13.24 (1.92) | 12.70 (1.63) | -3.01 (12.41) |  | 20.84 (1.52) | 18.71 (1.84) | -10.19 (6.18) |  |
|  | 90th Percent | 22.54 (1.46) | 19.73 (0.66) | -12.24 (3.69) |  | 30.42 (2.06) | 35.14 (3.14) | 15.46 (6.19) |  |
| 5. Abnormals,<br>No Warnings,<br>Non-experts | Mean | 7.57 (0.75) | 12.20 (0.82) | 61.87 (10.89) | <0.01* | 9.47 (0.97) | 15.76 (0.93) | 67.55 (12.48) | <0.05* |
|  | Median | 2.77 (0.13) | 3.77 (0.34) | 36.29 (12.19) |  | 3.43 (0.23) | 4.75 (0.61) | 38.72 (14.83) |  |
|  | 75th Percent | 9.56 (1.62) | 17.41 (0.80) | 86.49 (26.43) |  | 11.94 (2.32) | 20.21 (1.06) | 75.02 (30.43) |  |
|  | 90th Percent | 20.66 (2.37) | 35.48 (3.70) | 74.37 (29.63) |  | 26.21 (2.40) | 48.12 (2.90) | 85.12 (19.76) |  |

## References

[1] A. Dibden, J. Offman, D. Parmar, J. Jenkins, J. Slater, K. Binysh, J. McSorley, S. Scorfield, P. Cumming, X-H Liao, M. Ryan, D. Harker, G. Stevens, N. Rogers, R. Blanks, S. Sellars, J. Patnick, and S W Duffy. Reduction in interval cancer rates following the introduction of two-view mammography in the UK breast screening programme. British Journal of Cancer, 110(3):560–564, feb 2014.

[2] A. K. Hackshaw, N. J. Wald, M. J. Michell, S. Field, and A. R. Wilson. An investigation into why two-view mammography is better than one-view in breast cancer screening. Clinical Radiology, 55(6):454–458, 2000.

[3] Dipti Gupta and Sarah M Friedewald. Lesion localization using digital breast tomosynthesis: Where did i go wrong? Journal of Breast Imaging, 1(2):143–150, 05 2019.

[4] P. Skaane, A. I. Bandos, R. Gullien, E. B. Eben, U. Ekseth, and U. Haakenaasen. Comparison of digital mammography alone and digital mammography plus tomosynthesis in a population-based screening program. Radiology, 267(1):47–56, 2013.

[5] K. Lang, I. Andersson, A. Rosso, A. Tingberg, P. Timberg, and S. Zackrisson. Performance of one-view breast tomosynthesis as a stand-alone breast cancer screening modality: results from the malmö breast tomosynthesis screening trial, a population-based study. European Radiology, 26(1):184–190, 2016.

[6] George J. W. Partridge, Iain Darker, Jonathan J. James, Keshthra Satchithananda, Nisha Sharma, Alexandra Valencia, William Teh, Humaira Khan, Elizabeth Muscat, Michael J. Michell, and Yan Chen. How long does it take to read a mammogram? Investigating the reading time of digital breast tomosynthesis and digital mammography. European Journal of Radiology, 177:111535, 2024.

[7] D. Bernardi, P. Macaskill, M. Pellegrini, M. Valentini, C. Fantò, L. Ostillio, and N. Houssami. Breast cancer screening with tomosynthesis (3d mammography) with acquired or synthetic 2d mammography compared with 2d mammography alone (storm-2): a population-based prospective study. The Lancet Oncology, 20(3):360–372, 2019.

[8] Veronica Magni, Andrea Cozzi, Simone Schiaffino, Anna Colarieti, and Francesco Sardanelli. Artificial intelligence for digital breast tomosynthesis: Impact on diagnostic performance, reading times, and workload in the era of personalized screening. European Journal of Radiology, 158:110631, 2023.

[9] Sarah E. Hickman, Nicholas R. Payne, Richard T. Black, Yuan Huang, Andrew N. Priest, Sue Hudson, Bahman Kasmai, Arne Juette, Muzna Nanaa, and Fiona J. Gilbert. Deep learning algorithms for breast cancer detection in a uk creening cohort: As stand-alone readers and combined with human readers. Radiology, 313(2):e233147, 2024. PMID: 39560480.

[10] I. Sechopoulos. A review of breast tomosynthesis: Part i. the image acquisition process. Medical Physics, 40(1):014301, 2013.

[11] S. Vedantham, A. Karellas, G. R. Vijayaraghavan, and D. B. Kopans. Digital breast tomosynthesis: state of the art. Radiology, 277(3):663–684, 2015.

[12] Dominik Resch, Roberto Lo Gullo, Jonas Teuwen, Florian Semturs, Johannes Hummel, Andreas Resch, and Katja Pinker. Ai-enhanced mammography with digital breast tomosynthesis for breast cancer detection: Clinical value and comparison with human performance. Radiology: Imaging Cancer, 6(4):e230149, 2024.

[13] R.S. Lewis, Sunshine J.H., and Bhargavan M. A portrait of breast imaging specialists and of the interpretation of mammography in the united states. American Journal of Roentgenology, 187(5):W456– W468, 2006.

[14] Rickard Strandberg, Marike Gabrielson, Kamila Czene, Per Hall, and Keith Humphreys. Estimating distributions of breast cancer onset and growth in a swedish mammography screening cohort. *Cancer Epidemiology*, Biomarkers & Prevention, 32(5):801–808, 2023.

[15] Mark D Halling-Brown, Lucy M Warren, Dominic Ward, Emma Lewis, Alistair Mackenzie, Matthew G Wallis, Louise Wilkinson, Rosalind M Given-Wilson, Rita McAvinchey, and Kenneth C Young. Optimam mammography image database: a large scale resource of mammography images and clinical data, 2020.

[16] U.S. Food and Drug Administration. Viewfinder software version 1.1, k223501, 2023. FDA 510(k) Clearance Date: April 21, 2023.

[17] Diana L. Miglioretti, Charlotte C. Gard, Patricia A. Carney, Tracy L. Onega, Diana S. M. Buist, Edward A. Sickles, Karla Kerlikowske, Robert D. Rosenberg, Bonnie C. Yankaskas, Berta M. Geller, and Joann G. Elmore. When radiologists perform best: the learning curve in screening mammogram interpretation. Radiology, 253(3):632–640, 2009.

[18] H. Zhu and B. E. Doğan. American joint committee on cancer’s staging system for breast cancer, eighth edition: Summary for clinicians. European Journal of Breast Health, 17(3):234–238, 2021.

[19] Cancer Research UK. Stages and grades of breast cancer. Cancer Research UK, 2024. Accessed November 26, 2024.

[20] Mahesh K Shetty. Presurgical localization of breast abnormalities: an overview and analysis of 202 cases. Indian Journal of Surgical Oncology, 1(4):278–283, 2010.

[21] James Chiu, Davide Bova, Georgia Spear, Jacob Ecanow, Alyssa Choate, Pierre Besson, and Calin Caluser. Improving lesion location reproducibility in handheld breast ultrasound. Diagnostics, 14(15):1602, 2024.

[22] Nicolas Roduit. Weasis dicom viewer. Accessed 5 April 2023, 2023. Version 4.1.0.

[23] Winston Haynes et al. Wilcoxon rank sum test. Encyclopedia of systems biology, 3(1):2354–2355, 2013.

